# Systematic analysis of off-label and off-guideline cancer therapy usage in a real-world cohort of 153,122 U.S. patients

**DOI:** 10.1101/2023.01.17.23284689

**Authors:** Ruishan Liu, Lisa Wang, Shemra Rizzo, Marius Rene Garmhausen, Navdeep Pal, Sarah Waliany, Sarah McGough, Yvonne G. Lin, Zhi Huang, Joel Neal, Ryan Copping, James Zou

**Author notes:** Equal contributions (Lisa Wang, Shemra Rizzo, Marius Rene Garmhausen, Navdeep Pal, Sarah Waliany). Senior authors (Ryan Copping, James Zou).

## Abstract

Patients with cancer may be given treatments that are not officially approved (off-label) or recommended by guidelines (off-guideline) for multiple reasons including a lack of effective approved treatments. Here we present a systematic characterization of the patterns of off-label and off-guideline usage in 153,122 U.S. patients with 14 common cancer types using a large electronic health record (EHR)-derived de-identified database. We find that 18.3% and 3.9% of patients have received at least one line of off-label and off-guideline cancer drugs, respectively. Out of the 14 malignancies investigated, advanced bladder cancer has the highest proportion with 8.1% of patients receiving off-guideline treatments, most of which are recommended for non-small cell lung cancer. Patients with worse performance status, in later lines, or treated at academic hospitals are significantly more likely to receive off-label and off-guideline drugs. Underrepresented minority patients are less likely to receive off-guideline treatments in several cancer types. To quantify how predictable off-guideline usage is, we developed machine learning models to predict which drug a patient is likely to receive based on their clinical characteristics and previous treatments. Finally, we demonstrate that our systematic analysis of large real-world cohorts can identify interesting candidates for potential label expansion by identifying off-label treatments that demonstrate effectiveness in the real world setting. For example, we find that hormonal agents approved for breast cancer are used off-label in patients with ovarian cancer. Moreover, these hormonal agents show promising effectiveness in ovarian cancer with adjusted hazard ratio 0.53 (0.44, 0.65) compared to standard-of-care. This work demonstrates the power of large-scale computational analysis of real-world data for investigating non-standard cancer treatment usages.

## Introduction

In modern oncology care, anticancer drugs are frequently used beyond official regulatory approval (off-label) or outside of curated evidence-based guidelines (off-guideline)^1^. Off-label drug use in the U.S. is defined as prescribing medicines inconsistent with the label approved by the U.S. Food and Drug Administration (FDA). In addition to FDA labels, the U.S. National Comprehensive Cancer Network has developed the Clinical Practice Guidelines in Oncology (NCCN Guidelines®) which describe recommendations for oncology care and are updated regularly. The guidelines often include, in addition to approved drugs, off-label drug usage that is supported by clinical evidence. This allows patients to benefit from the therapies months or years before FDA approval^2^. We refer to drug uses outside NCCN guideline recommendations as “off-guideline.”

Off-label use in cancer care is widespread and acknowledged by the National Cancer Institute (NCI) and the FDA^3,4^. A literature review of patients from different countries revealed that, as of 2016, 13% to 71% of adult patients with cancer received at least one off-label chemotherapy, and that off-label drug use outside standard treatment guidelines was between 7% and 31%^1,5^. Unfortunately, despite the widespread practice of treating patients with unapproved drugs, the clinical outcomes and adverse events associated with off-label or off-guideline therapy usage are not systematically collected or shared^6^. Moreover, there has been little systematic analysis of off-guideline usage in the U.S.

The use of drugs for an unlicensed indication does not necessarily imply a safety hazard. Approved labels are often limited in scope^5^ due to restrictive clinical trial populations^7^. There are many instances where use outside of the label is uncontroversial and known to be advantageous to the patients. For example, oxaliplatin which is approved in colorectal cancer is often used off-label and off-guideline in triple-negative breast cancer patients^8–10^ and carboplatin, which is not labeled for children, is routinely used for the treatment of solid tumors in pediatric patients^11^. Off-label usage enables treating patients diagnosed with specific tumor types where treatment options are limited and allows physicians to use their clinical judgment to determine suitable treatment for an individual patient. For example, in bladder cancer, carboplatin can be more conveniently administered than cisplatin^5^. Furthermore, once sufficient scientific evidence (e.g. publications showing treatment efficacy and patient safety) is given, approved drugs for one indication may be included in the guidelines for other indications.

Despite the widespread practice of off-label use, the lack of systematic assessment of usage patterns and clinical benefits remains a major concern^12^. Real-world data is starting to be used to address the lack of high quality data needed to characterize the prevalence and clinical outcomes of off-label and off-guideline use. Recent studies have illustrated the power of real-world data in assessing the prevalence, effectiveness and/or safety of off-label drugs in primary central nervous system lymphoma^13^, breast cancer^14,15^ and melanoma^16^. The FDA has also increasingly adopted real-world data to support FDA approval for oncology drugs and label expansions^17^. For example, the CDK4/6 inhibitor palbociclib was first approved in 2015 to treat women with estrogen receptor–positive, HER2-negative breast cancer. In 2019, the FDA expanded the palbociclib indications to include men on the basis of retrospective outcomes analysis of real-world data derived from EHRs^18^.

However, existing studies leveraging real-world data to investigate non-standard cancer treatment usage have several shortcomings. They have only been performed on small sample sizes, a single cancer, or a single drug, and they do not distinguish between off-label and off-guideline usage^14,19,20^. With the advancement of anticancer therapies and the increase of approved drugs, we can expect a continuous increase in the use of off-label and off-guidance drugs over time, particularly in novel combinations^21^. Hence, a better understanding of off-label and off-guidance usage is critical. To the authors’ knowledge, the present study is the first large-scale real world data analysis to fill this gap and address previous studies’ limitations. Specifically, we systematically analyze the prevalence of off-label and off-guidance drugs across 14 cancer types in 153,122 patients from 280 cancer clinics across the U.S. We further perform a temporal analysis to highlight trends of off-guidance usage over time as well as an assessment of patient’s characteristics that are associated with off-guidance usage. Finally, we provide three use cases to showcase the impactful benefit of large-scale systematic analyses of real-world data in the assessment of off-label and off-guidance drug usage.

## Results

### Summary of data and methodology

In our study, we investigate off-label and off-guideline usage of cancer drugs in 153,122 patients in the U.S. This study used the nationwide Flatiron Health electronic health record (EHR)-derived de-identified database. The Flatiron Health (FH) database is a longitudinal database, comprising de-identified patient-level structured and unstructured data, curated via technology-enabled abstraction^22,23^ (see Supplementary Table 1 for the summary statistics of patient characteristics). The de-identified data originated from approximately 280 cancer clinics (∼800 sites of care) and have been shown to be representative of patients with cancer in the U.S^22^. The study included patients diagnosed with exactly one of the following 14 cancers: advanced non-small cell lung cancer (aNSCLC) (n=48,853), metastatic colorectal cancer (mCRC) (n=22,838), metastatic breast cancer (mBC) (n=10,687), multiple myeloma (MM) (n=10,910), chronic lymphocytic leukemia (CLL) (n=9,389), metastatic pancreatic cancer (mPCa) (n=8,414), ovarian cancer (OC) (n=5,220), adult acute myeloid leukemia (AML) (n=5,876), small cell lung cancer (SCLC) (n=6,550), advanced bladder cancer (aBCa) (n=5,774), diffuse large B-cell lymphoma (DLBCL) (n=6,308), advanced melanoma (aMel) (n=5,763), follicular lymphoma (FL) (n=3,318) and hepatocellular carcinoma (HCC) (n=3,222). We removed patients diagnosed with multiple cancers from all of the downstream analyses. The data included treatments with 241 antineoplastic drugs (Supplementary Table 2). For the analysis of predictive biomarkers, we used the nationwide (U.S.-based) de-identified Flatiron Health-Foundation Medicine clinico-genomic database (FH-FMI CGDB)

We retrieved the FDA approved drug list for each cancer type from the records of the National Cancer Institute (NCI) at the National Institutes of Health (NIH) up to June 20, 2022 (Supplementary Table 3)^24^. We use a conservative definition of off-label and off-guideline usage. For each cancer, a drug that is not approved under any condition (e.g. not approved in any dosage) is considered off-label. For example, a drug’s approved indication may be quite specific with regard to the patient subgroup, prior therapies received or if the drug is given as monotherapy or in combination with another drug. For our analyses, treatments received not meeting these detailed conditions were not counted as off-label usage. The main factor defining off-label use in our analysis was the tumor type. In addition, we retrieved the recommended drug list of NCCN Guidelines for each cancer type from the NCCN Compendia (Supplementary Table 4). The NCCN Guidelines are recognized as the standard of care for oncology treatment. For each cancer, a drug that is not recommended under any condition by NCCN Guidelines is considered off-guideline.

### Overview of off-label and off-guideline usage patterns

Treatment in accordance with NCCN Guidelines is generally regarded as “appropriate” and usually covered by health insurance, even when it is outside of FDA labeling. Thus, we differentiate between off-label use that is and is not included in the NCCN Guidelines. For example, carboplatin does not have official FDA approval for mBC, SCLC, aBCa, DLBCL, aMel and FL, but is recommended by NCCN Guidelines for those malignancies and commonly administered in clinical practice. In the FH database, 18.3% patients were treated with at least one off-label drug and 3.9% of patients were treated with at least one off-guideline drug; see Supplementary Table 5 and Supplementary Table 6 for the statistics of off-label and off-guideline usage in different lines of treatments for each cancer type.

Figure 2 summarizes the cross-use of FDA approved and NCCN recommended drugs between different cancer types in the FH database. The most commonly administered off-label treatments were carboplatin, etoposide and bendamustine. The most commonly used off-guideline therapies include leuprolide, pemetrexed and bevacizumab. We found that the most prevalent off-label usage is in SCLC, with borrowed FDA approved drugs from aNSCLC and OC — 77.6%, 23.2% and 8.2% of patients with SCLC received carboplatin, cisplatin and paclitaxel, respectively. The most prevalent off-guideline usage is in aBCa, with borrowed NCCN recommended drugs from aNSCLC and OC — 4.1% and 1.5% of patients with aBCa received pemetrexed and paclitaxel protein-bound, respectively.

**Figure 1.**
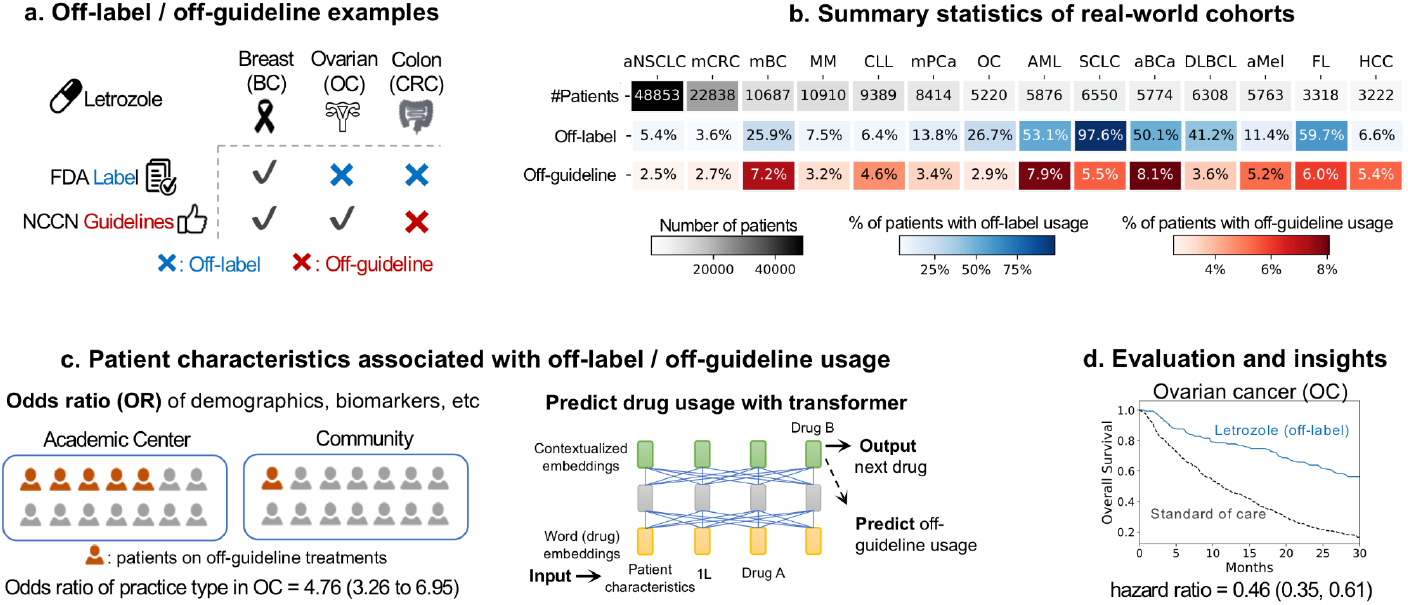
Real-world data analysis workflow and applications. **(a)** For each cancer, we identify drug usage outside of FDA approval as “off-label” and drug usage not in accordance with NCCN Guidelines as “off-guideline”. **(b)** The total number of patients and the proportion of patients with off-label usage and off-guideline usage for 14 cancers in Flatiron Health EHR-derived database. **(c)** We characterize how a patient’s characteristics are associated with off-guideline usage. Adjusted odds ratio analysis was used to estimate the extent in which patient characteristics were associated with off-guideline usage. We use a transformer-based language model to predict the treatment trajectory of patients and off-guideline usage. **(d)** We evaluate the effectiveness of off-guideline drugs using survival analysis with propensity score weighting.

**Figure 2.**
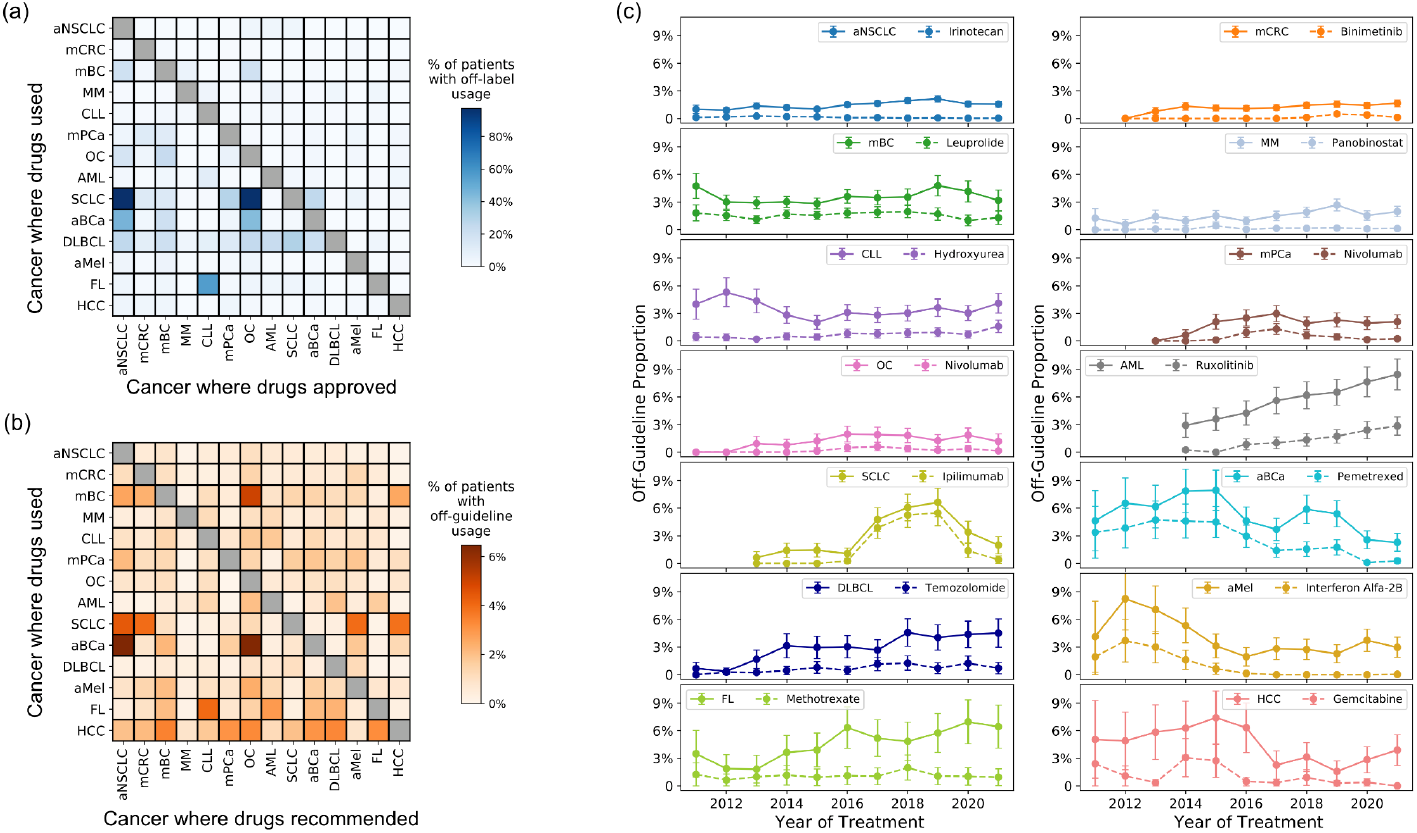
The cross-use of (**a**) approved and (**b**) recommended drugs for 14 cancer types. The (i, j) grid shows the proportion of patients with cancer i (row i) receiving drugs that are (a) off-label or (b) off-guideline, but (a) approved or (b) recommended by NCCN Guidelines in the cancer j (column j). (**c**) Temporal trend of off-guideline usage. For each cancer, we plot the proportion of patients receiving any off-guideline treatment (solid line) and the commonly administered off-guideline drug (dashed line) for each year.

### Temporal trends

We analyzed the temporal trends of patients receiving off-guideline treatments (Figure 2) and off-label treatments (Supplementary Figure 1) in each year in the FH database. We focus on the off-guideline analysis in the main text due to the fact that off-guideline drugs recommended by NCCN Guidelines are generally also regarded as appropriate and are covered by health insurance. Off-guideline usage has been increasing in recent years for mCRC, AML, DLBCL, FL and HCC, due to the increasing use of off-guideline targeted therapies such as binimetinib and vemurafenib for mCRC and ruxolitinib for AML. Binimetinib was approved by the FDA in 2018 to treat BRAF mutation-positive melanoma. Consistent with this, 94% of the patients with mCRC who received binimetinib and tested for BRAF status in FH tested positive for BRAF mutations. While ruxolitinib is not in the treatment guideline for AML, it has shown promising results in patients with AML in early clinical trials^25^. In contrast, the off-guideline usage has decreased for mBC, CLL and aBCa. For example, the off-guideline usage in aBCa dropped from 10% in 2015 to 2% in 2020, mainly due to the approval of the immunotherapy pembrolizumab. After its initial FDA approval in 2014 (for melanoma patients), pembrolizumab was rapidly used for patients with aBCa (Figure 2). Off-guideline drug use for aBCa (e.g. pemetrexed) especially declined after pembrolizumab was approved for aBCa by the FDA in 2017.

### Characteristics of patients who received off-guideline treatments

We next investigate which patient characteristics are associated with increased usage of off-guideline treatments in the FH database. For each cancer, we fit a logistic regression model with patients characteristics to predict whether or not they received off-guideline drugs (Methods). We summarized the odds ratio of different patients characteristics associated with off-guideline usage in Figure 3 and off-label usage in Supplementary Figure 2. Several clinical characteristics of patients are significantly associated with increased off-guideline usage. Across the 14 cancer types, patients in later lines of treatment and with worse performance status (higher ECOG values) are more likely to receive off-guideline treatments. This suggests that patients who are more advanced in their cancer progression, and thus have fewer options, are more likely to be given off-guideline drugs as treating physicians try to identify effective treatment options. Patients treated at academic hospitals are also more likely to receive off-guideline drugs compared to patients treated at community clinics (median adjusted OR = 2.17 across 14 cancer types). Patients with tumors that are positive for specific biomarkers such as EGFR and KRAS in the FH database tended to have less off-guideline usage, perhaps due to the availability of FDA approved targeted therapies for patients whose tumors harbor these mutations.

**Figure 3.**
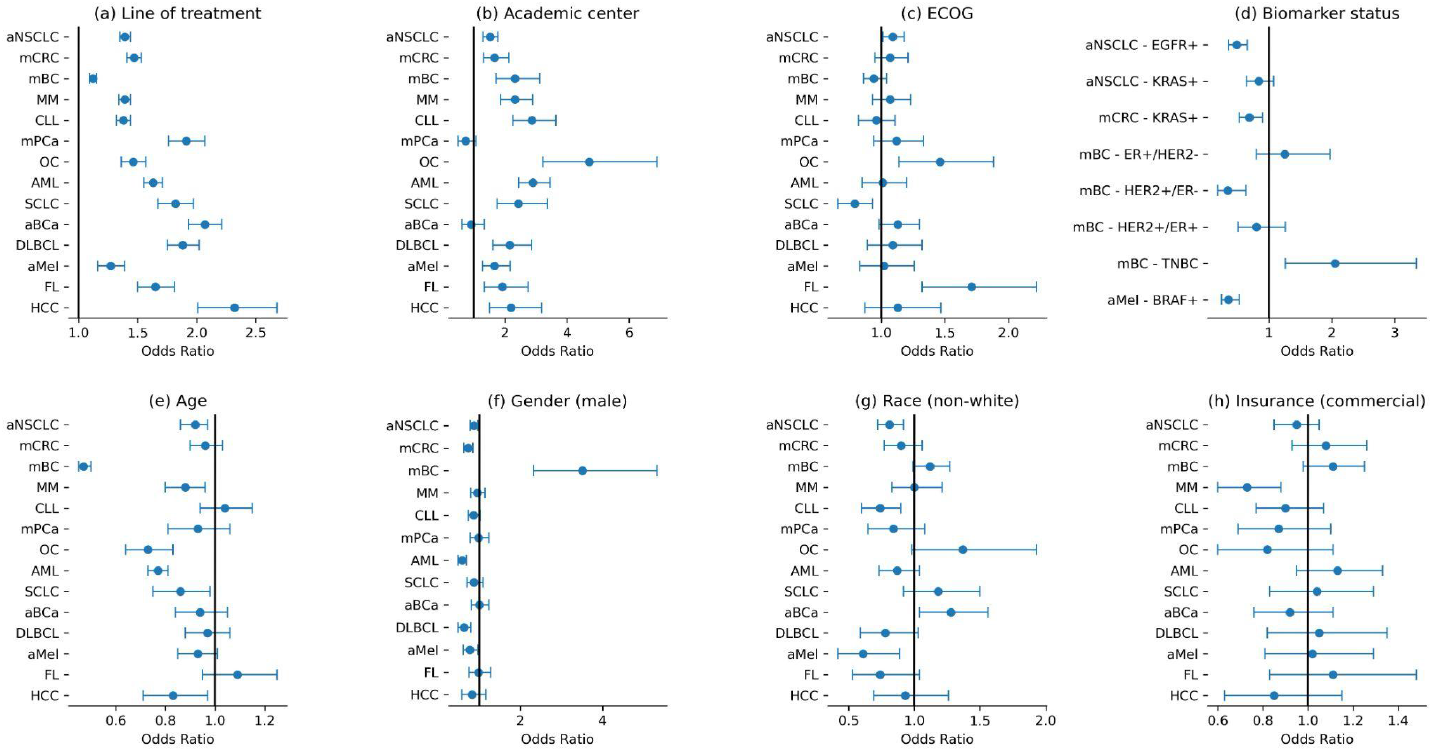
Adjusted odds ratio for off-guideline drug usage for (a) line of treatment, (b) practice type (academic or community center), (c) ECOG value, (d) biomarker status, (e) age in increments of 10 years, (f) sex (male or not), (g) race (non-white or not), (h) insurance category (commercial or not, on the patient’s first insurance record). Here the odds ratios are adjusted for age, sex, race, ECOG, staging, practice type, insurance type, year of receiving treatment, histology (for patients with aNSCLC and OC), smoking status (for patients with aNSCLC and SCLC) and biomarkers status. Error bars correspond to 95% binomial confidence intervals.

Certain patient demographics were also associated with off-guideline usage. Interestingly, younger patients receive more off-guideline usage. One possible explanation is that clinicians may feel that younger patients can better tolerate off-guideline treatment, which may have more uncertain side effects. Self-reported underrepresented minorities (non-white) are less likely to receive off-guideline treatments in several cancer types (aMel, aNSCLC, CLL, DLBCL and AML), potentially reflecting historically well-documented racial disparities in cancer care (Figure 3). Such disparities impact the minority patient care experience in myriad ways that may affect off-label and off-guideline use, including reduced access to novel or experimental therapies^26^, reduced access to specialist care and resources^27^, and lack of information and communication about treatment options^28–30^. There is no broad trend between off-guideline usage and insurance type (e.g. Medicare, Medicaid and commercial insurance), which is also considered a proxy for socioeconomic status. There is no consistent association between sex and off-guideline usage except for mBC. For mBC, less than 1% of patients are male and they are significantly more likely to receive off-guideline treatments^31^. Because male breast cancer patients are rare, few are included in clinical trials, and recommendations on the clinical management of male breast cancer patients are extrapolated from the population of female breast cancer patients in trials. Generally, although males are treated similarly to females, there are some differences. For example, aromatase inhibitors for mBC are given concurrently with gonadotropin-releasing hormone analogs (e.g., leuprolide) for males, but without those analogs for most women, especially post-menopausal women, according to the NCCN Guidelines for breast cancer.

It is interesting to quantify how well factors captured in the EHR can holistically predict when a patient might receive off-guideline treatments. When off-guideline usage is predictable, models could potentially alert clinicians for additional treatment options to consider. It is also useful to know when off-guideline usage is difficult to predict, as this quantifies the extent to which factors or randomness beyond the guidelines and standard clinical features influence treatment recommendations. To quantify off-guideline predictability, we developed several machine learning algorithms to model the trajectory of patient treatments. Overall, a sophisticated language-model type transformer neural network achieved the best performance in predicting which patient would receive off-guideline treatments in a particular line. The transformer is first trained with self-supervised learning to predict which drug a patient receives in each line based on the patient’s baseline characteristics (e.g. age, ECOG, staging, etc.) and previous lines of treatments (Supplementary Table 7). The patient embedding from the transformer is then trained to predict off-guideline usage, where the model achieves AUPRC of 0.12, 0.20 and 0.28 for first, second and third lines of therapy, respectively (Supplementary Table 8). One reason that the AUPRC is relatively low is because the prediction task is heavily imbalanced, with only 3.9% of patients receiving off-guideline treatment (Supplementary Table 6). The relative proportion of off-guideline usage increases for later lines, consistent with the increase in AUPRC. The modest prediction performance of state-of-the-art machine learning models also suggests that off-guideline usage decisions are often affected by factors outside of what is typically captured in the EHR or the guidelines. Interestingly, HCC and AML are the most predictable cancer types, with AUPRC ranging from 0.15 to 0.75 across the lines, suggesting more patterned off-guideline usage.

Systematic real-world data analysis of off-label and off-guideline usage can generate valuable insights such as potential targets for label expansion, adoption patterns of new therapies, and effectiveness of new drug combinations. We next conduct case studies to illustrate these three applications.

### Real-world data (RWD) indicate the benefit of hormone therapy in ovarian cancer

The role of hormone therapy in treating ovarian cancer has been proposed in the NCCN guidelines. Recommended uses have generally been relegated to the less common epithelial ovarian cancers such as grade 1 endometrioid and low-grade serous carcinomas for which hormonal therapy is a recommended therapy in the adjuvant and/or maintenance setting. Furthermore, the efficacy of letrozole as an adjuvant monotherapy in comparison to paclitaxel/carboplatin plus letrozole is currently under investigation^32^. Patients deemed to be too frail or ill for cytotoxic chemotherapies are sometimes given hormonal therapies, which also has the benefit of not inducing chemo-resistance.

We evaluated the off-label uses of hormone therapy in OC using the FH database. Hormone therapies are mostly used in 2nd and later lines of treatment for ovarian cancer, where they were associated with better survival compared to non-hormone therapies in the FH database (hormone drugs are listed in Supplementary Table 9), with adjusted HR of 0.56, 0.63 and 0.65 for letrozole, tamoxifen and anastrozole, respectively (Table 1). Similar results were also observed in additional analyses of patients who received these same three hormonal therapies as single agents (i.e. monotherapy as opposed to combination therapy) in 2nd and later lines (Supplementary table 10). Overall, 15.4% of ovarian cancer patients receive hormone therapy in 2nd or later lines. After the adjustments of propensity weighting, the two comparative cohorts (patients receiving hormone therapy vs. patients receiving non-hormone therapy) were balanced based on all patient characteristics including demographics, disease conditions and treatment dates (Supplementary Figure 3). Additionally, we identified TP53 as a potential predictive biomarker using the FH-FMI CGDB database. Patients with any TP53 mutation were observed to have worse survival when treated with letrozole (adjusted interaction HR=2.32), tamoxifen (adjusted interaction HR=1.80) or anastrozole (adjusted interaction HR=1.66) compared to TP53 wild-type patients receiving the same treatment.

**Table 1.**
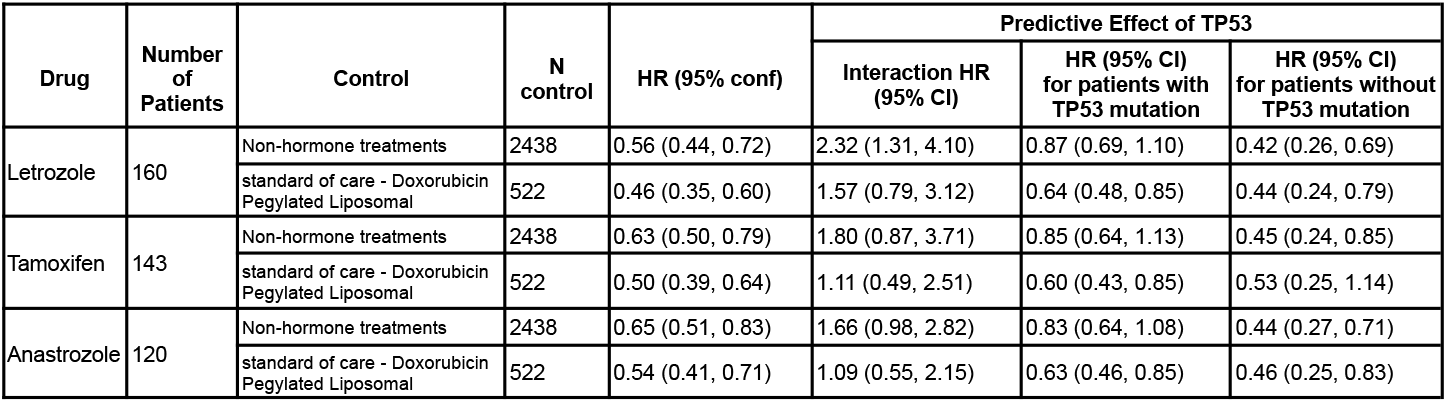
Evaluation of hormone therapy in ovarian cancer for 2L+. Here the experiment arm consists of patients who received treatments containing the hormone therapies (e.g. letrozole alone or in combination with other drugs). The control arm includes patients 1) who received treatments not containing any hormone agents; 2) standard of care, defined as the most commonly used treatment. Hazard ratios are adjusted for age, sex, race, ECOG, stage, histology and molecular alterations, practice type, insurance type, year of receiving treatment.

| Drug | Number of Patients | Control | N control | HR (95% conf) | Predictive Effect of TP53 |  |  |
| --- | --- | --- | --- | --- | --- | --- | --- |
|  |  |  |  |  | Interaction HR (95% CI) | HR (95% CI) for patients with TP53 mutation | HR (95% CI) for patients without TP53 mutation |
| Letrozole | 160 | Non-hormone treatments | 2438 | 0.56 (0.44, 0.72) | 2.32 (1.31, 4.10) | 0.87 (0.69, 1.10) | 0.42 (0.26, 0.69) |
|  |  | standard of care - Doxorubicin Pegylated Liposomal | 522 | 0.46 (0.35, 0.60) | 1.57 (0.79, 3.12) | 0.64 (0.48, 0.85) | 0.44 (0.24, 0.79) |
| Tamoxifen | 143 | Non-hormone treatments | 2438 | 0.63 (0.50, 0.79) | 1.80 (0.87, 3.71) | 0.85 (0.64, 1.13) | 0.45 (0.24, 0.85) |
|  |  | standard of care - Doxorubicin Pegylated Liposomal | 522 | 0.50 (0.39, 0.64) | 1.11 (0.49, 2.51) | 0.60 (0.43, 0.85) | 0.53 (0.25, 1.14) |
| Anastrozole | 120 | Non-hormone treatments | 2438 | 0.65 (0.51, 0.83) | 1.66 (0.98, 2.82) | 0.83 (0.64, 1.08) | 0.44 (0.27, 0.71) |
|  |  | standard of care - Doxorubicin Pegylated Liposomal | 522 | 0.54 (0.41, 0.71) | 1.09 (0.55, 2.15) | 0.63 (0.46, 0.85) | 0.46 (0.25, 0.83) |

### Early adoption of immunotherapy

There is growing interest in measuring the adoption patterns of new immunotherapies, but relatively little is known about the frequency of immunotherapy usage *before* FDA approval^33^. As a case study, we report the FDA approval history of nivolumab for different cancer types in Table 2. Nivolumab was first approved in aMel in 2014, and its off-label use in other cancer types has been observed since then. For example, nivolumab was not approved for SCLC until August 2018, but 8.3% of patients with SCLC already received nivolumab before this date. As of the writing of this paper, nivolumab is not yet approved for mPCa and we evaluate its off-label use in 2+ lines in the FH database in Supp. Table 11. Across different cancer types, we find that a non-negligible fraction of patients received nivolumab in non-approved tumor types before official FDA approval, suggesting enthusiasm for new immunotherapy adoption.

**Table 2.** FDA approval history for nivolumab. For each cancer type, we report the date when nivolumab is first approved by the FDA, and the number of patients that received nivolumab before and after the approval date in the FH database.

| Cancer | Approval Date | Before Approval |  |  | After Approval |  |  |
| --- | --- | --- | --- | --- | --- | --- | --- |
|  |  | Number of Patients on Nivolumab | Total Number of Patients | % of Patients on Nivolumab | Number of Patients on Nivolumab | Total Number of Patients | % of Patients on Nivolumab |
| aMel | 12/22/2014 | 1 | 1114 | 0.1% | 2911 | 4866 | 59.8% |
| aNSCLC | 03/04/2015 | 3 | 14449 | 0.0% | 7734 | 36938 | 20.9% |
| mCRC | 08/01/2017 | 53 | 10836 | 0.5% | 287 | 14541 | 2.0% |
| HCC | 09/22/2017 | 58 | 1545 | 3.8% | 603 | 1802 | 33.5% |
| SCLC | 08/16/2018 | 342 | 4128 | 8.3% | 308 | 2751 | 11.2% |
| aBCa | 08/19/2021 | 354 | 5456 | 6.5% | 47 | 554 | 8.5% |
| mPCa | Unapproved | 71 | 8414 | 0.8% | - | - | - |

### RWD provides evidence suggesting ineffective immunotherapy combination, consistent with clinical trials

Ipilimumab and nivolumab are both immune checkpoint inhibitors that have complementary mechanisms of action. Nivolumab was approved for SCLC in 2018; mounting evidence illustrated that the addition of ipilimumab to nivolumab was more effective than nivolumab monotherapy for NSCLC^34^, but until recently the efficacy of the combination for SCLC was unclear. In our study of the FH database, we find that patients with SCLC who received nivolumab and ipilimumab (N=214) did not show improved overall survival compared to patients who received only nivolumab (N=381) in 2+ lines with adjusted HR of 1.03 (95% CI (0.85, 1.25). Consistent with our findings on real-world patients, recent clinical trials for SCLC reported that overall survival was similar between the group treated with nivolumab alone and the group treated with nivolumab plus ipilimumab^35,36^, and the nivolumab plus ipilimumab combination was removed from SCLC guideline in 2021. This shows how real-world data analysis could generate hypotheses for clinical trials and potential guideline changes.

## Discussion

Our systematic analysis of 14 common cancer types in 153,122 U.S. patients demonstrates that prescribing a drug outside its official labeling and practice guidelines is relatively widespread in oncology. To the best of our knowledge, this is the largest study investigating off-label and off-guideline usage of anticancer drugs using real-world data. As more and higher quality real-world data become available, our computational framework could be expanded beyond the Flatiron Health data. Real-world data is an important resource for label expansion because it reflects how patients respond to diverse treatments in the real-world setting. In this work, as a case study, we identify that some patients with ovarian cancer may benefit from hormone therapies that are approved for breast cancer but are currently off-label in ovarian cancer.

We found that certain patient characteristics were associated with increased likelihood of receiving off-guideline treatments: younger age, academic settings, and the receipt of later lines of therapy. Additionally, our results suggest that underrepresented minorities are less likely to receive off-guideline treatments. Few studies have explored racial disparities in the context of off-label usage, in particular in the context of cancer; however, one psychiatry study found that minority patients were less likely to fill off-label prescriptions than whites when restricted to private insurance^37^. Still, further research and more high-quality data are needed to fully explore the dynamics at play and whether these racial disparities in off-label and off-guideline treatment imply racial disparities in quality of patient care.

There are good reasons for some cancer treatments to be used off-label or off-guideline. For example, many chemotherapies are older drugs that received approval in one cancer indication many years ago, but manufacturers of those drugs may lack incentives to conduct the additional clinical trials needed to pursue official FDA approval for other tumor types^24^. Moreover, cytotoxic chemotherapies have the same overall mechanism of stopping fast reproducing cancer cells, so subclasses of these chemotherapies are sometimes used interchangeably by oncologists even if only one drug received regulatory approval (e.g. cisplatin and carboplatin are both platinum-based therapies). In addition, cancer treatments are often given in combination (e.g., combination of chemotherapies such as FOLFOX and FOLFIRI for mCRC). Since it is challenging for the FDA to try to approve all chemotherapy combinations, a combination chemotherapy regimen could contain one or more chemotherapies that are not approved for the tumor type being treated^3^. In addition, our machine learning analysis shows that off-guideline usage has low predictability, suggesting that complex factors beyond treatment guidelines and clinical information commonly captured in the medical records may influence decisions to use off-guideline therapies.

There are several limitations to be further explored in the future. Although we used the state-of-the-art propensity adjustment methods to mitigate selection bias, there could be unmeasured confounders in real-world data (e.g. tumor grade for ovarian cancer patients). In addition, detailed safety analyses were challenging due to the sparsity of related endpoints in the Flatiron Health database. Moreover, we focus our analysis on drugs captured in the FH database. While we excluded patients who were known to have more than one malignancy, it is possible that some patients who were included in the analysis had multiple malignancies but with the diagnosis code for the other malignancy not available in the EMR. To mitigate some of these missing data challenges in this work, we used a strict definition of off-label or off-guideline drug — a drug that is not approved or recommended under any condition for a specific cancer type. The official labels and practice guidelines may also include specific requirements and conditions, such as usage in a special subpopulation, in a specified dosage and through a specific administration route. Analysis of drug usage not in accordance with detailed approval and recommendation conditions is an important future direction.

Identifying opportunities where access to approved therapies can potentially be broadened to benefit patients with other cancer types and stages is an important area of focus for drug developers. This paper demonstrates that large-scale analysis of high quality real world data can help to characterize the landscape of off-label and off-guideline treatment usage and identify potential label expansion opportunities for further research. This kind of analysis has the potential to provide rapid and quantitative insights into the effectiveness of off-label and off-guideline treatments that can then be used to inform research and development efforts.

## Data Availability

Requests for data sharing of these de-identified data by license/by permission for the specific purpose of replicating results in this manuscript can be submitted to.

## Methods

### Flatiron Health dataset

The data that support the findings of this study come from the nationwide EHR-derived de-identified Flatiron Health database (March 2022 datacut). The Flatiron Health database is a longitudinal database, comprising de-identified patient-level structured and unstructured data, curated via technology-enabled abstraction^22,23^. The Flatiron Health data is considered one of the industry’s leading research databases in oncology due to the rigorous data curation and abstraction processes as well as publications demonstrating their efforts to validate outcomes. In previous validation studies comparing the Flatiron Health mortality data to the gold standard National Death Index, the sensitivity of mortality capture in an aNSCLC population was shown to be 91%, and the impact of remaining missing deaths on survival analyses was minimal^38–40^. In addition to curation accuracy, the Flatiron Health data is harmonized and aggregated across approximately 280 cancer clinics across the country, which enables its data to be more representative than single healthcare EHR. The majority of patients in the database originate from community oncology settings; relative community/academic proportions may vary depending on study cohort. Data provided to investigators was de-identified and subject to obligations to prevent re-identification and protect patients’ confidentiality. The data that support the findings of this study originated by Flatiron Health, Inc. and Foundation Medicine, Inc. Requests for data sharing of these de-identified data by license/by permission for the specific purpose of replicating results in this manuscript can be submitted to. Portions of research conducted by Flatiron Health were approved with waiver of informed consent by the WCG Institutional Review Board prior to study conduct.

### FH-FMI CGDB data

The Flatiron Health dataset is linked with the comprehensive genomic profiling results from Foundation Medicine (FMI) as the de-identified Flatiron Health-Foundation Medicine clinico-genomic database (FH-FMI CGDB, datacut March 31, 2022). We used the FH-FMI CGDB dataset to identify potential genetic biomarkers for hormone therapy in OC. Retrospective longitudinal clinical data were derived from electronic health record (EHR) data, comprising patient-level structured and unstructured data, curated via technology-enabled abstraction, and were linked to genomic data derived from FMI comprehensive genomic profiling (CGP) tests in the FH-FMI CGDB by de-identified, deterministic matching^41^. The FH and FMI data match was conducted by an independent third party^41^. Data provided to investigators were de-identified and subject to obligations to prevent re-identification and protect patients’ confidentiality. Portions of research conducted by Flatiron Health were approved with waiver of informed consent by the WCG Institutional Review Board prior to study conduct.

Genomic alterations were identified via comprehensive genomic profiling (CGP) of more than 300 cancer-related genes on FMI’s next-generation sequencing (NGS) tests^42–44^. Eligible patients must receive the Foundation Medicine test on a tumor sample with pathologist-confirmed histology that is consistent with Flatiron Health abstracted tumor type, and their Foundation Medicine specimen collection date must be no earlier than 30 days after the initial diagnosis date in Flatiron Health. All the patients have at least two documented clinical visits in the Flatiron network on or after January 1, 2011. This study included 3,253 patients diagnosed with OC (removed patients diagnosed with multiple cancers) who underwent FMI CGP testing.

### Identification of off-label drugs

The data included treatments with 241 antineoplastic drugs (Supplementary Table 2). The drug biosimilars are grouped together (Supplementary Table 12). The FDA approved drug list for each cancer type was retrieved from the records of the National Cancer Institute (NCI) at the National Institutes of Health (NIH) (data up to June 20, 2022) (Supplementary Table 3)^24^. We use a conservative definition of off-label usage. For each cancer, a drug that is not approved under any condition is considered off-label.

### Identification of off-guideline drugs

The recommended drug list of NCCN Guidelines for each cancer type was retrieved from the NCCN Compendia (Supplementary Table 4). The NCCN Guidelines are recognized as the standard for clinical direction. Similar to the conservative definition of off-label usage, a drug that is not recommended under any condition for a given cancer type by NCCN Guidelines (October 2022 version) is considered off-guideline.

### Removal of patients with multiple cancers

For rigorous off-label usage analysis, we focused on patients with only one cancer. We filtered patients in two steps. First, we reviewed the diagnosis records for all patients and removed those with ICD diagnosis codes in multiple cancers. Apart from the 14 cancers in our analysis, we also checked for and removed patients with diagnosis codes for chronic myelogenous leukemia (CML), acute lymphocytic leukemia (ALL), cutaneous T-cell lymphomas (CTCLs), myelofibrosis (MF), renal cell carcinoma (RCC), head and neck cancer, gastric cancer, malignant neoplasm of brain, prostate cancer, and endometrial cancer. Second, we removed all the patients who qualified as patients in multiple Flatiron datasets for different cancer types.

### Odds ratio evaluating associations between patient characteristics and off-label usage

The adjusted odds ratio (aOR) between each covariate and off-label usage is defined as the exponential of the logistic regression coefficient adjusted for baseline confounders. The categorical variables are encoded as dummy variables and continuous variables are standardized. For each covariate, we filter out patients with missing values. We control for the following baseline confounders: age, sex, race, ECOG, staging, year of receiving treatment, practice type, insurance type in the patient’s first insurance record, histology (aNSCLC and OC), smoking status (aNSCLC and SCLC) and biomarkers status (Supplementary Table 13). When different lines of treatments are analyzed together, we also include line number as a confounder. The ECOG value and biomarker status are selected as the closest value assessed within a -180 to +7 days window around the start date of the line. All the baseline confounders are one-hot encoded for categories with at least 5% of patients.

### Quantifying the predictability of off-guideline drug usage

In order to quantify the predictability of off-guideline drug usage, we trained several machine learning models to predict which patients would receive off-guideline treatment in each line. Here we focus on off-guideline rather than off-label, because off-guideline treatments are clinically more interesting and are also more challenging to predict. We interpret the best performance of the machine learning models as approximating the predictability of off-guideline usage. The best performing model overall is a transformer-based language model that we developed to model the trajectory of patient treatments^45^. This model considers drugs and patient characteristics as individual tokens. For example, the drug “carboplatin” and sex “male” are fed to the language model as tokens. We define the end of each line, death and censoring as special tokens. The task of the language model is to predict which drug is likely to follow a sequence of tokens; that is, to predict the probability of a patient receiving a given drug considering their characteristics and all of the prior treatments. For each line of treatment, the drugs are sequenced alphabetically. The sequence of tokens are passed to an embedding layer with dimension of 200 and a positional encoding layer with dropout rate of 0.2. A square attention mask is used to mask out all the tokens in the future positions. We use two layers of transformer encoder with two attention heads and feedforward network with dimension of 200. The output of the transformer (learned embeddings) is passed to a linear layer with log softmax. We train the model for 100 epochs and use a batch size of 20 and stochastic gradient descent with a learning rate starting from 5 and decaying 5% every epoch.

To evaluate the performance of the transformer in predicting which drugs a patient takes in each line, we input the patient characteristics and sequence of treatments prior to a given line to the model. The drug within the given line is predicted iteratively until the prediction of end of the line, death or censoring. The accuracy of predicting the drug combinations in each line is defined as

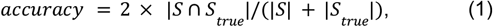

where *S*(*S*_*true*_) is the learned (true) set of drugs in the given line of treatment.

To predict the off-guideline usage in each line, we input the patient embedding learned by the transformer to a logistic regression model. We compare the transformer’s performance with a logistic regression model trained on the original patient characteristics. Other machine learning models, such as random forest, trained on patient characteristics have similar performance as logistic regression. In the experiments, we randomly split the patients into training, validation and test set, with ratio 70%, 15% and 15%, respectively. We fit the model on the training set, use the validation set for early stopping, and report the model’s performance on the test set.

### Performance of off-label drugs

To evaluate the performance of one off-label drug for one cancer type, we compare the survival behavior of two groups — the experiment group with all the patients on treatments containing that drug (ie. drug given alone or in combination with another drug treatment) and the control group with all the patients not receiving that drug. We used Cox proportional-hazards model to assess hazard ratio and its 95% confidence interval. To perform survival analysis, we set the index date (time zero) as the start date of the corresponding line of treatment. Patients were followed until death for real world overall survival (OS) analysis. The censoring dates are chosen as their last visit dates to the clinics.

To obtain unbiased estimates, we used inverse probability of treatment weighting (IPTW) to adjust for baseline confounding factors. In the survival analysis, patient *i* is weighted by ω^(*i*)^ in Equation 2, where *e*^(*i*)^ is the propensity score for patient *i* and *Z*^(*i*)^ is an indicator to represent whether patient *i* is in the experiment group or not. For example, when analyzing the prognostic effect of gene mutation on overall survival, patients with a mutated gene of interest compose the experiment group (*Z*^(*i*)^ =1) and patients without mutations in that gene are in the control group (*Z*^(*i*)^ =0). The propensity score *e*^(*i*)^ is estimated by a logistic regression model in Equation 3, where 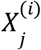 indicates the *j*-th baseline covariate for patient *i*. The *n*_*X*_confounders are effectively balanced between the experiment and control groups after adjusting by propensity score.

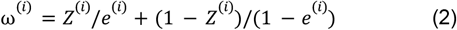

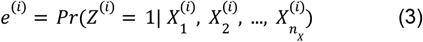

The baseline confounders *X* is the same as used in odds ratio analysis.

### Ovarian cancer analysis

For ovarian cancer patients in the Flatiron Health dataset, 2L therapy represents the 2nd regimen received after the patient’s initial ovarian cancer diagnosis. This differs from the traditional definition of line of therapy, where LoT is specifically 1st treatment regimen, 2nd treatment regimen, etc that a patient receives after their advanced or metastatic diagnosis.

### Predictive biomarker identification

The interactive Cox proportional hazards model between one gene mutation and one treatment includes gene mutation status *G*, treatment status *T*, a gene-by-treatment interaction term *G* × *T* and the baseline confounders *X*_*j*_, as defined in Equation 4.

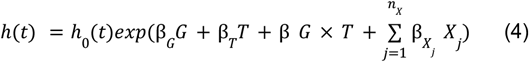

Here *h*(*t*) is the hazard function at time *t, h*_0_(*t*) is the baseline hazard. The antilog of the regression coefficient *exp*(β_*G*_), *exp*(β_*T*_), *exp*(β) and 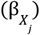 produce hazard ratios for the corresponding variables. A gene-by-treatment interaction HR *exp*(β) larger than 1 indicates that the treatment effect is worse in patients whose tumors harbor a mutation in that gene, compared to patients without that tumor genetic mutation. Here we use the same set of confounders *X* as in the odds ratio analysis. To avoid the immortal time bias that may arise from death before getting the chance to take genomic profiling tests^44^, we penalized the Cox proportional hazards model with left-truncation method with the dates when patients receive the FMI tests^47^.

## Acknowledgements

We thank G. Sledge Jr., M. Schuessler, C. Harbron, S. Maund, M. Taylor, M. Ballinger, and V. Steffen for comments and discussions. L.W., S.R., M.R.G, N.P., S.M., Y.G.L., and R.C. are supported by funding from Roche.

## Author contributions

R.L., L.W., S.R., M.R.G, N.P., S.W., J.N., R.C. and J.Z. designed the project. R.L. developed methodology and conducted analysis with assistance from all the authors. S.M., Y.G.L., Z.H., S.W., J.N. provided clinical interpretations. All the authors contributed to paper writing. R.C. and J.Z. supervised the project.

## Data and code availability

The data used in this study were licensed from Flatiron Health (https://flatiron.com/real-world-evidence/) and Foundation Medicine. These de-identified data may be made available upon request; interested researchers can contact. The open source Python code for the computational analysis is available at https://github.com/RuishanLiu/off-label.

## Supplementary Figures and Tables

**Supplementary Figure 1.**
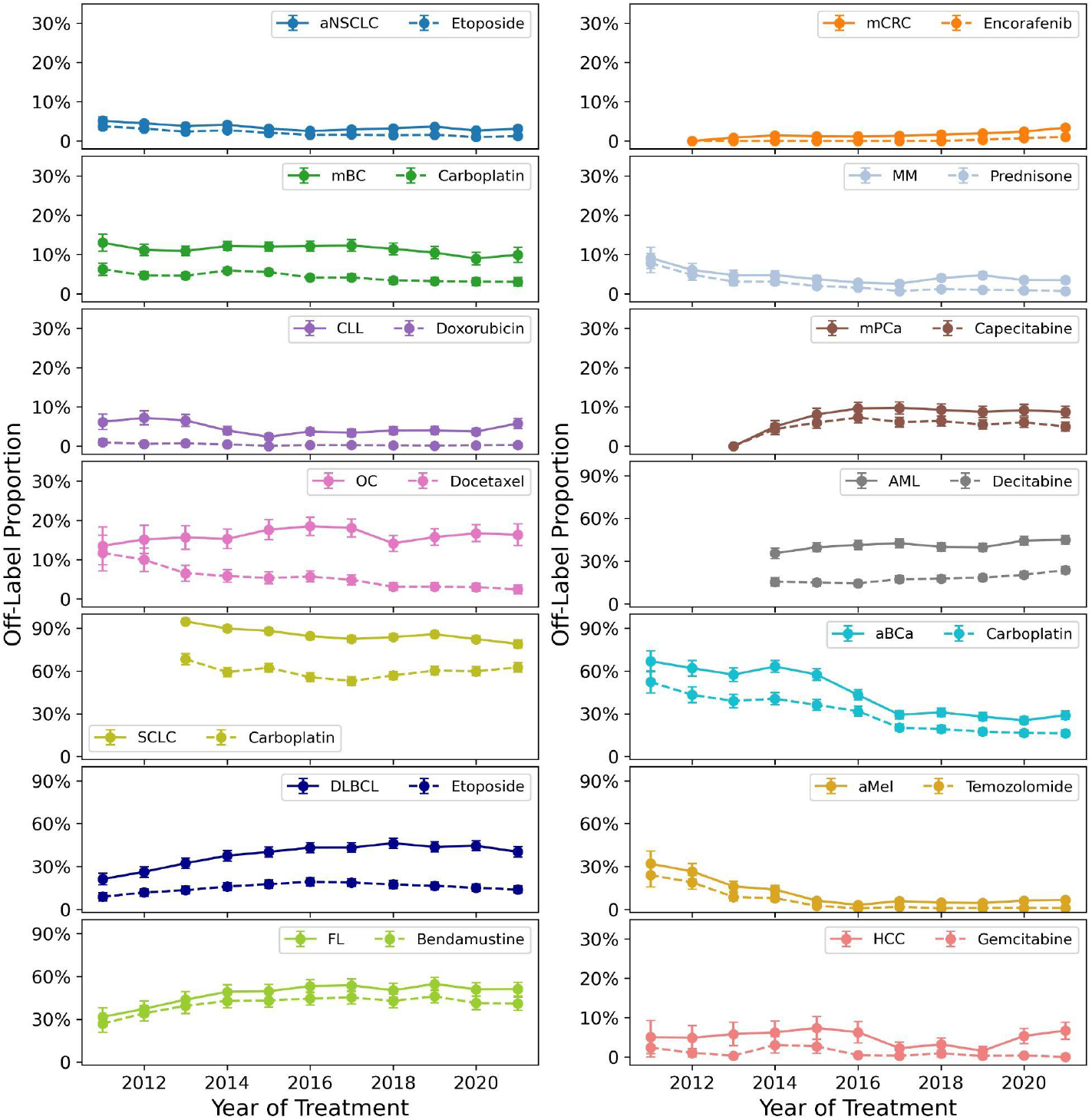
Temporal trend of off-label usage. For each cancer, we plot the proportion of patients receiving any off-label treatment (solid line) and the most commonly administered off-label drug (dashed line) each year.

**Supplementary Figure 2.**
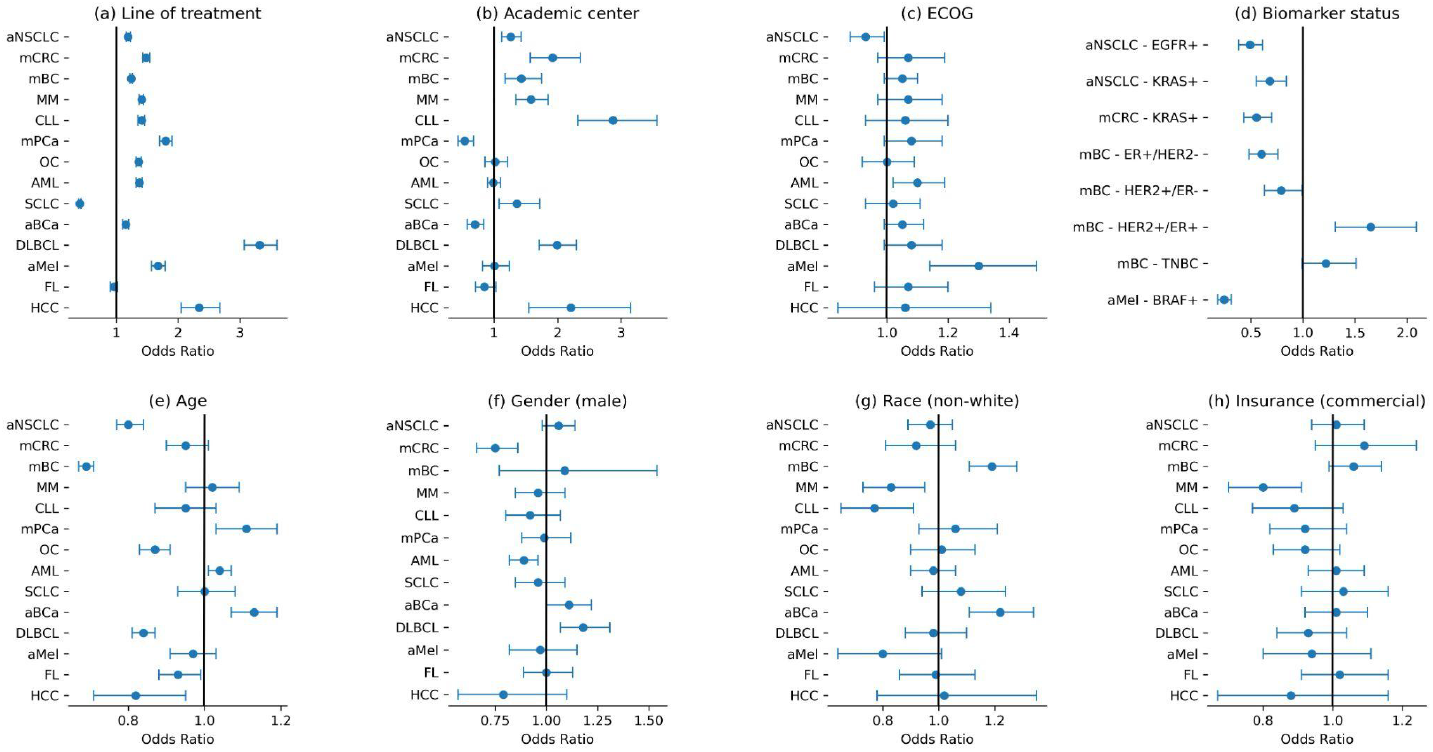
Adjusted odds ratio for off-label drug usage for (a) line of treatment, (b) practice type (academic center or not), (c) ECOG value, (d) biomarker status, (e) age in increments of 10 years, (f) sex (male or not), (g) race (non-white or not), (h) insurance category (commercial or not). Here the odds ratio is adjusted for age, sex, race, ECOG, staging, practice type, insurance type, year of receiving treatment, histology (aNSCLC and OC), smoking status (aNSCLC and SCLC) and biomarkers status.

**Supplementary Figure 3.**
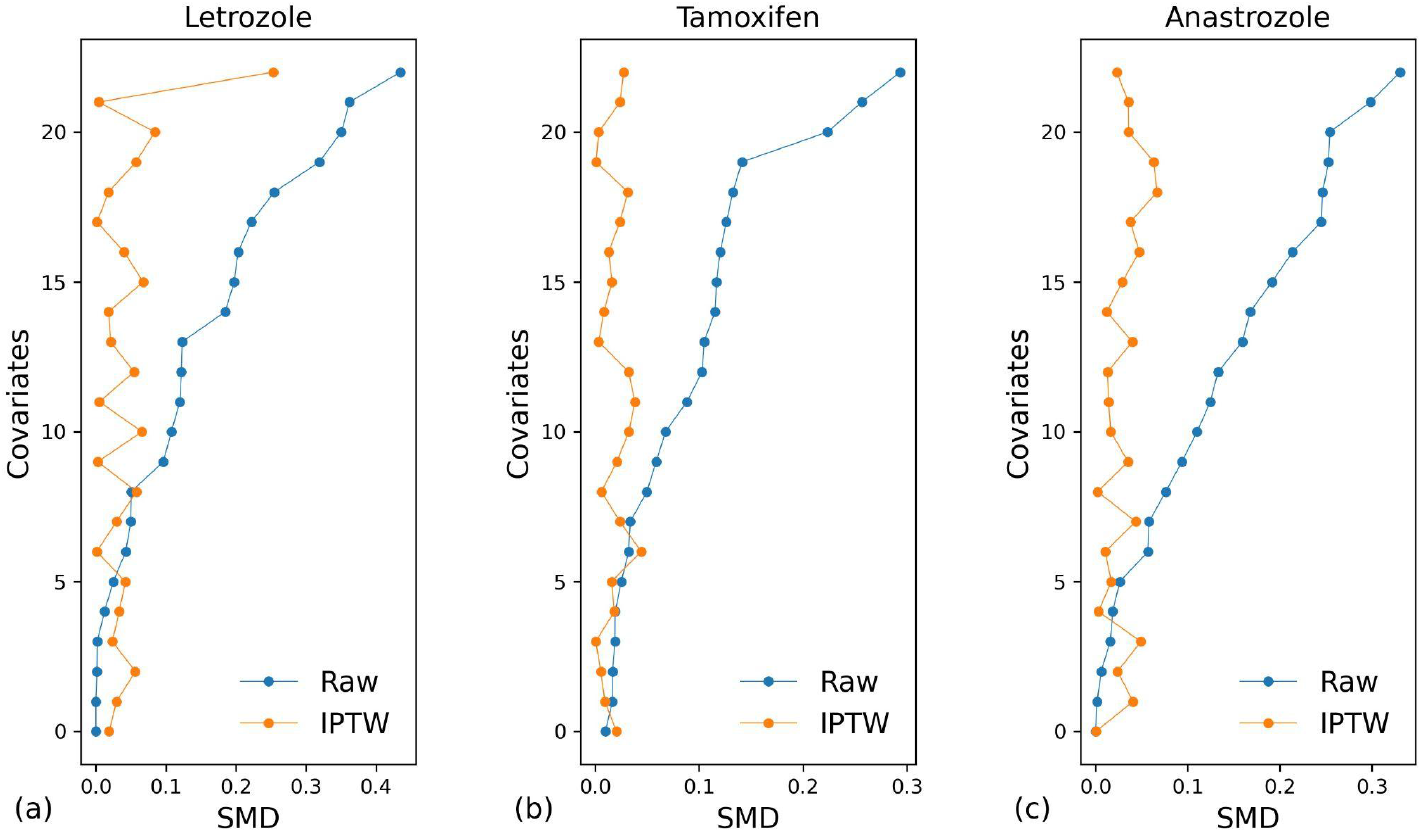
Balance assessment between experiment and control cohorts for baseline covariates. Here we use the comparison of hormone therapy with standard of care on OC for 2L+ as an example. We plot the standardized mean difference (SMD) for every baseline covariate between the experiment cohort (e.g. patients who received letrozole) and control cohort (e.g. patients who received doxorubicin pegylated liposomal). SMD close to 0 represents that the two cohorts are balanced. The inverse propensity weighting (IPTW) used in our analysis effectively balances the two cohorts. Raw corresponds to the unadjusted cohorts.

**Supplementary Table 1.** Summary statistics of 14 common cancer types in the Flatiron Health database.

|  | aNSCLC | mCRC | mBC | MM | CLL | mPCa | OC | AML | SCLC | aBCa | DLBCL | aMel | FL | HCC |
| --- | --- | --- | --- | --- | --- | --- | --- | --- | --- | --- | --- | --- | --- | --- |
| **Total** | 48853 | 22838 | 10687 | 10910 | 9389 | 8414 | 5220 | 5876 | 6550 | 5774 | 6308 | 5763 | 3318 | 3222 |
| **Age: Mean (IQR)** |  |  |  |  |  |  |  |  |  |  |  |  |  |  |
| **Median (IQR)** | 69 (61 - 75) | 64 (55 - 73) | 64 (55 - 73) | 69 (61 - 76) | 71 (63 - 77) | 68 (61 - 75) | 66 (57 - 74) | 68 (56 - 77) | 68 (61 - 74) | 73 (65 - 78) | 68 (57 - 75) | 67 (57 - 76) | 66 (56 - 74) | 66 (60 - 73) |
| **Sex - Female** |  |  |  |  |  |  |  |  |  |  |  |  |  |  |
| **F** | 23265 (47.6%) | 10072 (44.1%) | 10567 (98.9%) | 5014 (46.0%) | 3554 (37.9%) | 3821 (45.4%) | 5220 (100.0%) | 2465 (42.0%) | 3390 (51.8%) | 1549 (26.8%) | 2840 (45.0%) | 1872 (32.5%) | 1668 (50.3%) | 627 (19.5%) |
| **ECOG** |  |  |  |  |  |  |  |  |  |  |  |  |  |  |
| **0** | 9437 (19.3%) | 6426 (28.1%) | 2224 (20.8%) | 2019 (18.5%) | 2690 (28.7%) | 1947 (23.1%) | 1351 (25.9%) | 617 (10.5%) | 1189 (18.2%) | 1303 (22.6%) | 1223 (19.4%) | 1654 (28.7%) | 929 (28.0%) | 574 (17.8%) |
| **1** | 14473 (29.6%) | 5937 (26.0%) | 1759 (16.5%) | 2161 (19.8%) | 2129 (22.7%) | 2862 (34.0%) | 1241 (23.8%) | 964 (16.4%) | 1998 (30.5%) | 1713 (29.7%) | 1084 (17.2%) | 1297 (22.5%) | 567 (17.1%) | 826 (25.6%) |
| **2** | 5516 (11.3%) | 1640 (7.2%) | 642 (6.0%) | 858 (7.9%) | 480 (5.1%) | 884 (10.5%) | 358 (6.9%) | 429 (7.3%) | 877 (13.4%) | 645 (11.2%) | 360 (5.7%) | 423 (7.3%) | 130 (3.9%) | 347 (10.8%) |
| **3** | 1320 (2.7%) | 366 (1.6%) | 165 (1.5%) | 322 (3.0%) | 101 (1.1%) | 210 (2.5%) | 91 (1.7%) | 109 (1.9%) | 211 (3.2%) | 169 (2.9%) | 116 (1.8%) | 133 (2.3%) | 26 (0.8%) | 58 (1.8%) |
| **4** | 67 (0.1%) | 24 (0.1%) | 19 (0.2%) | 26 (0.2%) | 9 (0.1%) | 10 (0.1%) | 9 (0.2%) | 10 (0.2%) | 11 (0.2%) | 8 (0.1%) | 14 (0.2%) | 13 (0.2%) | 1 (0.0%) | 5 (0.2%) |
| **missing** | 18040 (36.9%) | 8445 (37.0%) | 5878 (55.0%) | 5524 (50.6%) | 3980 (42.4%) | 2501 (29.7%) | 2170 (41.6%) | 3747 (63.8%) | 2264 (34.6%) | 1936 (33.5%) | 3511 (55.7%) | 2243 (38.9%) | 1665 (50.2%) | 1412 (43.8%) |
| **Group Stage** |  |  |  |  |  |  |  |  |  |  |  |  |  |  |
| **0** | 3 (0.0%) | 3 (0.0%) | 2 (0.0%) | 0 (0.0%) | 0 (0.0%) | 0 (0.0%) | 0 (0.0%) | 0 (0.0%) | 0 (0.0%) | 21 (0.4%) | 0 (0.0%) | 21 (0.4%) | 0 (0.0%) | 0 (0.0%) |
| **I** | 3599 (7.4%) | 562 (2.5%) | 1300 (12.2%) | 10910 (100.0%) | 9389 (100.0%) | 394 (4.7%) | 571 (10.9%) | 5876 (100.0%) | 222 (3.4%) | 93 (1.6%) | 737 (11.7%) | 468 (8.1%) | 320 (9.6%) | 88 (2.7%) |
| **II** | 2246 (4.6%) | 2324 (10.2%) | 3223 (30.2%) | 0 (0.0%) | 0 (0.0%) | 1247 (14.8%) | 387 (7.4%) | 0 (0.0%) | 227 (3.5%) | 435 (7.5%) | 1081 (17.1%) | 940 (16.3%) | 470 (14.2%) | 145 (4.5%) |
| **III** | 10694 (21.9%) | 5297 (23.2%) | 2467 (23.1%) | 0 (0.0%) | 0 (0.0%) | 576 (6.8%) | 2289 (43.9%) | 0 (0.0%) | 1216 (18.6%) | 487 (8.4%) | 1198 (19.0%) | 1448 (25.1%) | 1056 (31.8%) | 343 (10.6%) |
| **IV** | 31136 (63.7%) | 13873 (60.7%) | 2469 (23.1%) | 0 (0.0%) | 0 (0.0%) | 5666 (67.3%) | 1296 (24.8%) | 0 (0.0%) | 3820 (58.3%) | 2113 (36.6%) | 1866 (29.6%) | 1790 (31.1%) | 1460 (44.0%) | 732 (22.7%) |
| **missing** | 1175 (2.4%) | 779 (3.4%) | 1226 (11.5%) | 0 (0.0%) | 0 (0.0%) | 531 (6.3%) | 677 (13.0%) | 0 (0.0%) | 1065 (16.3%) | 2625 (45.5%) | 1426 (22.6%) | 1096 (19.0%) | 12 (0.4%) | 1914 (59.4%) |
| **1L** |  |  |  |  |  |  |  |  |  |  |  |  |  |  |
| **All** | 48853 | 22838 | 10687 | 10910 | 9389 | 8414 | 5220 | 5876 | 6550 | 5774 | 6308 | 5763 | 3318 | 3222 |
| **Chemotherapy** | 22264 (45.6%) | 8226 (36.0%) | 2512 (23.5%) | 362 (3.3%) | 841 (9.0%) | 7945 (94.4%) | 3960 (75.9%) | 4246 (72.3%) | 5216 (79.6%) | 3944 (68.3%) | 139 (2.2%) | 205 (3.6%) | 32 (1.0%) | 42 (1.3%) |
| **Immunotherapy** | 13314 (27.3%) | 265 (1.2%) | 30 (0.3%) | 0 (0.0%) | 7 (0.1%) | 53 (0.6%) | 17 (0.3%) | 2 (0.0%) | 1216 (18.6%) | 1620 (28.1%) | 3 (0.0%) | 4090 (71.0%) | 1 (0.0%) | 268 (8.3%) |
| **Hormone_therapy** | 156 (0.3%) | 19 (0.1%) | 4967 (46.5%) | 3 (0.0%) | 13 (0.1%) | 3 (0.0%) | 145 (2.8%) | 74 (1.3%) | 5 (0.1%) | 5 (0.1%) | 1 (0.0%) | 22 (0.4%) | 3 (0.1%) | 2 (0.1%) |
| **Targeted_therapy** | 11504 (23.5%) | 13861 (60.7%) | 2879 (26.9%) | 9799 (89.8%) | 8268 (88.1%) | 68 (0.8%) | 909 (17.4%) | 1296 (22.1%) | 10 (0.2%) | 36 (0.6%) | 6115 (96.9%) | 1137 (19.7%) | 3238 (97.6%) | 2793 (86.7%) |
| **2L** |  |  |  |  |  |  |  |  |  |  |  |  |  |  |
| **All** | 20730 | 11771 | 7250 | 5943 | 4128 | 3183 | 2553 | 2789 | 2454 | 2433 | 1459 | 1830 | 878 | 858 |
| **Chemotherapy** | 7233 (34.9%) | 2317 (19.7%) | 1967 (27.1%) | 229 (3.9%) | 324 (7.8%) | 2861 (89.9%) | 1236 (48.4%) | 1699 (60.9%) | 1648 (67.2%) | 905 (37.2%) | 148 (10.1%) | 151 (8.3%) | 62 (7.1%) | 43 (5.0%) |
| **Immunotherapy** | 6934 (33.4%) | 237 (2.0%) | 25 (0.3%) | 2 (0.0%) | 11 (0.3%) | 61 (1.9%) | 17 (0.7%) | 2 (0.1%) | 673 (27.4%) | 1300 (53.4%) | 14 (1.0%) | 966 (52.8%) | 3 (0.3%) | 382 (44.5%) |
| **Hormone_therapy** | 48 (0.2%) | 13 (0.1%) | 2377 (32.8%) | 3 (0.1%) | 15 (0.4%) | 10 (0.3%) | 199 (7.8%) | 39 (1.4%) | 6 (0.2%) | 3 (0.1%) | 8 (0.5%) | 8 (0.4%) | 8 (0.9%) | 1 (0.1%) |
| **Targeted_therapy** | 5684 (27.4%) | 8945 (76.0%) | 2572 (35.5%) | 5349 (90.0%) | 3666 (88.8%) | 135 (4.2%) | 972 (38.1%) | 882 (31.6%) | 17 (0.7%) | 55 (2.3%) | 1251 (85.7%) | 609 (33.3%) | 779 (88.7%) | 400 (46.6%) |

**Supplementary Table 2.**
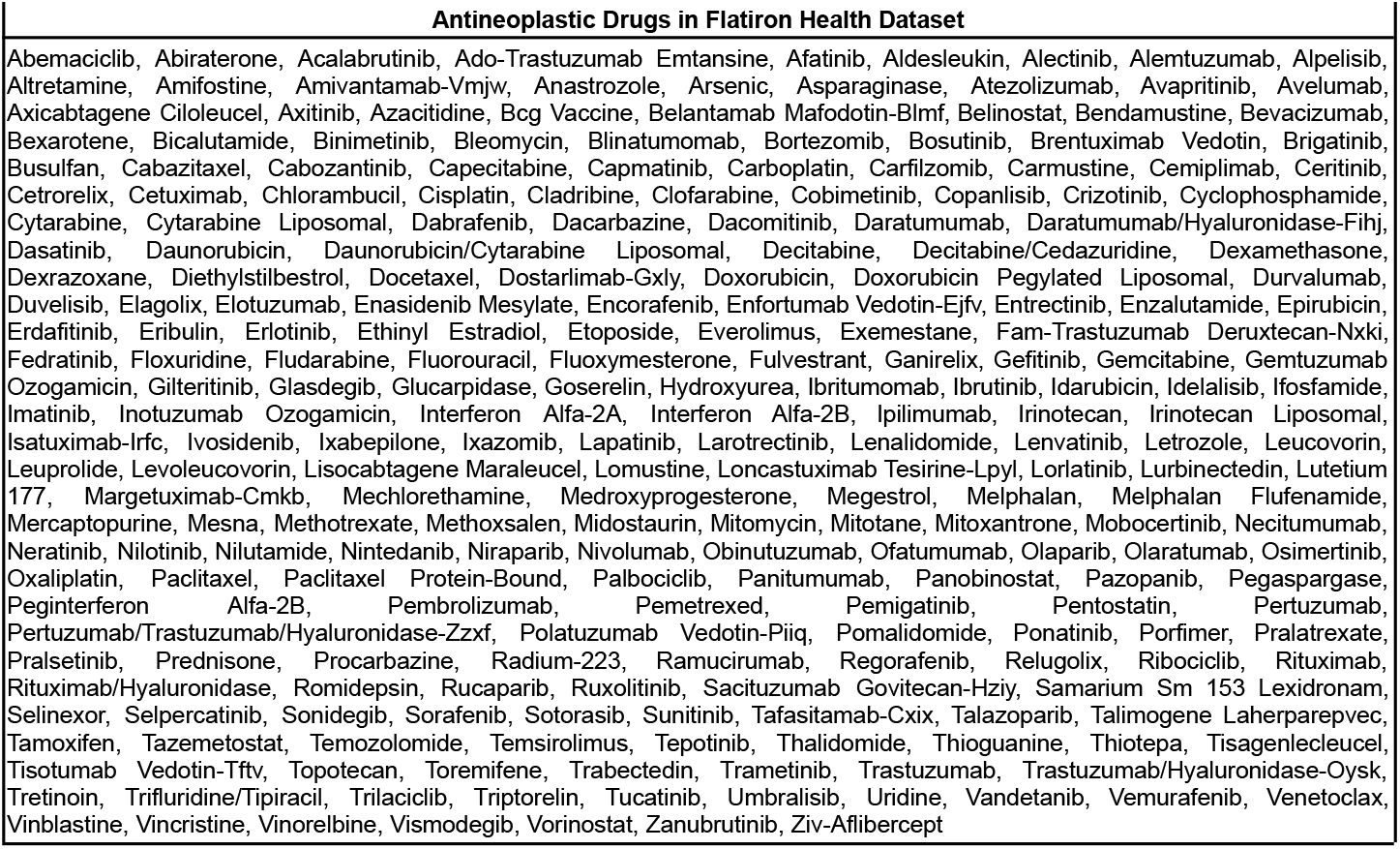
The 241 antineoplastic drugs included in the Flatiron Health dataset.

**Supplementary Table 3.** For each cancer, the FDA approved drug list from the records of the NCI up to June 20, 2022.

| Cancer | Drugs Approved by FDA |
| --- | --- |
| aNSCLC | Afatinib, Alectinib, Amivantamab-Vmjw, Atezolizumab, Bevacizumab, Brigatinib, Capmatinib, Cemiplimab, Ceritinib, Crizotinib, Dabrafenib, Dacomitinib, Docetaxel, Durvalumab, Entrectinib, Erlotinib, Everolimus, Gefitinib, Gemcitabine, Ipilimumab, Lorlatinib, Methotrexate, Mobocertinib, Necitumumab, Nivolumab, Osimertinib, Paclitaxel, Paclitaxel Protein-Bound, Pembrolizumab, Pemetrexed, Pralsetinib, Ramucirumab, Selpercatinib, Sotorasib, Trametinib, Tepotinib, Vinorelbine, Doxorubicin, Carboplatin, Paclitaxel, Gemcitabine, Cisplatin |
| mCRC | Bevacizumab, Irinotecan, Capecitabine, Cetuximab, Ramucirumab, Oxaliplatin, Fluorouracil, Ipilimumab, Pembrolizumab, Leucovorin, Trifluridine/Tipiracil, Nivolumab, Panitumumab, Regorafenib, Ziv-Aflibercept |
| mBC | Abemaciclib, Ado-Trastuzumab Emtansine, Alpelisib, Anastrozole, Atezolizumab, Capecitabine, Cyclophosphamide, Docetaxel, Doxorubicin, Epirubicin, Eribulin, Everolimus, Exemestane, Fam-Trastuzumab Deruxtecan-Nxki, Fluorouracil, Fulvestrant, Gemcitabine, Goserelin, Ixabepilone, Lapatinib, Letrozole, Margetuximab-Cmkb, Megestrol, Methotrexate, Neratinib, Olaparib, Paclitaxel, Paclitaxel Protein-Bound, Palbociclib, Pamidronate, Pembrolizumab, Pertuzumab, Pertuzumab/Trastuzumab/Hyaluronidase-Zzxf, Ribociclib, Sacituzumab Govitecan-Hziy, Talazoparib, Tamoxifen, Thiotepa, Toremifene, Trastuzumab, Trastuzumab/Hyaluronidase-Oysk, Tucatinib, Vinblastine |
| MM | Belantamab Mafodotin-Blmf, Bortezomib, Carfilzomib, Carmustine, Ciltacabtagene Autoleucel, Cyclophosphamide, Daratumumab, Daratumumab/Hyaluronidase-Fihj, Doxorubicin Pegylated Liposomal, Elotuzumab, Idecabtagene Vicleucel, Isatuximab-Irfc, Ixazomib, Lenalidomide, Melphalan, Melphalan Hydrochloride, Plerixafor, Pomalidomide, Selinexor, Thalidomide, Zoledronic Acid, Bortezomib, Doxorubicin, Dexamethasone |
| CLL | Acalabrutinib, Alemtuzumab, Bendamustine, Chlorambucil, Cyclophosphamide, Dexamethasone, Duvelisib, Fludarabine, Ibrutinib, Idelalisib, Obinutuzumab, Ofatumumab, Prednisone, Rituximab, Rituximab/Hyaluronidase, Venetoclax, Vincristine |
| mPCa | Paclitaxel Protein-Bound, Everolimus, Erlotinib, Fluorouracil, Gemcitabine, Irinotecan Liposomal, Olaparib, Mitomycin, Sunitinib, Leucovorin, Irinotecan, Oxaliplatin, Cisplatin |
| OC | Bevacizumab, Carboplatin, Cisplatin, Cyclophosphamide, Doxorubicin, Doxorubicin Pegylated Liposomal, Gemcitabine, Topotecan, Olaparib, Niraparib, Paclitaxel, Rucaparib, Thiotepa, Bleomycin, Etoposide, Vincristine, Vinblastine, Ifosfamide |
| AML | Arsenic, Azacitidine, Cyclophosphamide, Cytarabine, Daunorubicin, Dexamethasone, Doxorubicin, Enasidenib Mesylate, Gemtuzumab Ozogamicin, Gilteritinib, Glasdegib, Idarubicin, Ivosidenib, Midostaurin, Mitoxantrone, Prednisone, Thioguanine, Venetoclax, Vincristine, Tretinoin, Daunorubicin/Cytarabine Liposomal, Etoposide |
| SCLC | Atezolizumab, Doxorubicin, Durvalumab, Etoposide, Etoposide Phosphate, Everolimus, Lurbinectedin, Methotrexate, Nivolumab, Topotecan |
| aBCa | Atezolizumab, Cisplatin, Doxorubicin, Enfortumab Vedotin-Ejfv, Erdafitinib, Mitomycin, Nivolumab, Pembrolizumab, Sacituzumab Govitecan-Hziy, Thiotepa, Valrubicin, Gemcitabine, Methotrexate, Vinblastine |
| DLBCL | Axicabtagene Ciloleucel, Lisocabtagene Maraleucel, Loncastuximab Tesirine-Lpyl, Polatuzumab Vedotin-Piiq, Rituximab, Rituximab/Hyaluronidase, Selinexor, Tafasitamab-Cxix, Tisagenlecleucel, Rituximab, Cyclophosphamide, Doxorubicin, Vincristine, Prednisone |
| aMel | Aldesleukin, Binimetinib, Cobimetinib, Dabrafenib, Dacarbazine, Encorafenib, Ipilimumab, Nivolumab, Relatlimab-rmbw, Peginterferon Alfa-2B, Pembrolizumab, Interferon Alfa-2B, Talimogene Laherparepvec, Tebentafusp-tebn, Trametinib, Vemurafenib |
| FL | Axicabtagene Ciloleucel, Copanlisib, Ibritumomab, Lenalidomide, Lisocabtagene Maraleucel, Obinutuzumab, Interferon Alfa-2B, Rituximab, Rituximab/Hyaluronidase, Tazemetostat, Tisagenlecleucel, Rituximab, Cyclophosphamide, Doxorubicin, Vincristine, Prednisone |
| HCC | Atezolizumab, Bevacizumab, Cabozantinib, Infigratinib Phosphate, Lenvatinib, Nivolumab, Pembrolizumab, Pemigatinib, Ramucirumab, Regorafenib, Sorafenib |

**Supplementary Table 4.**
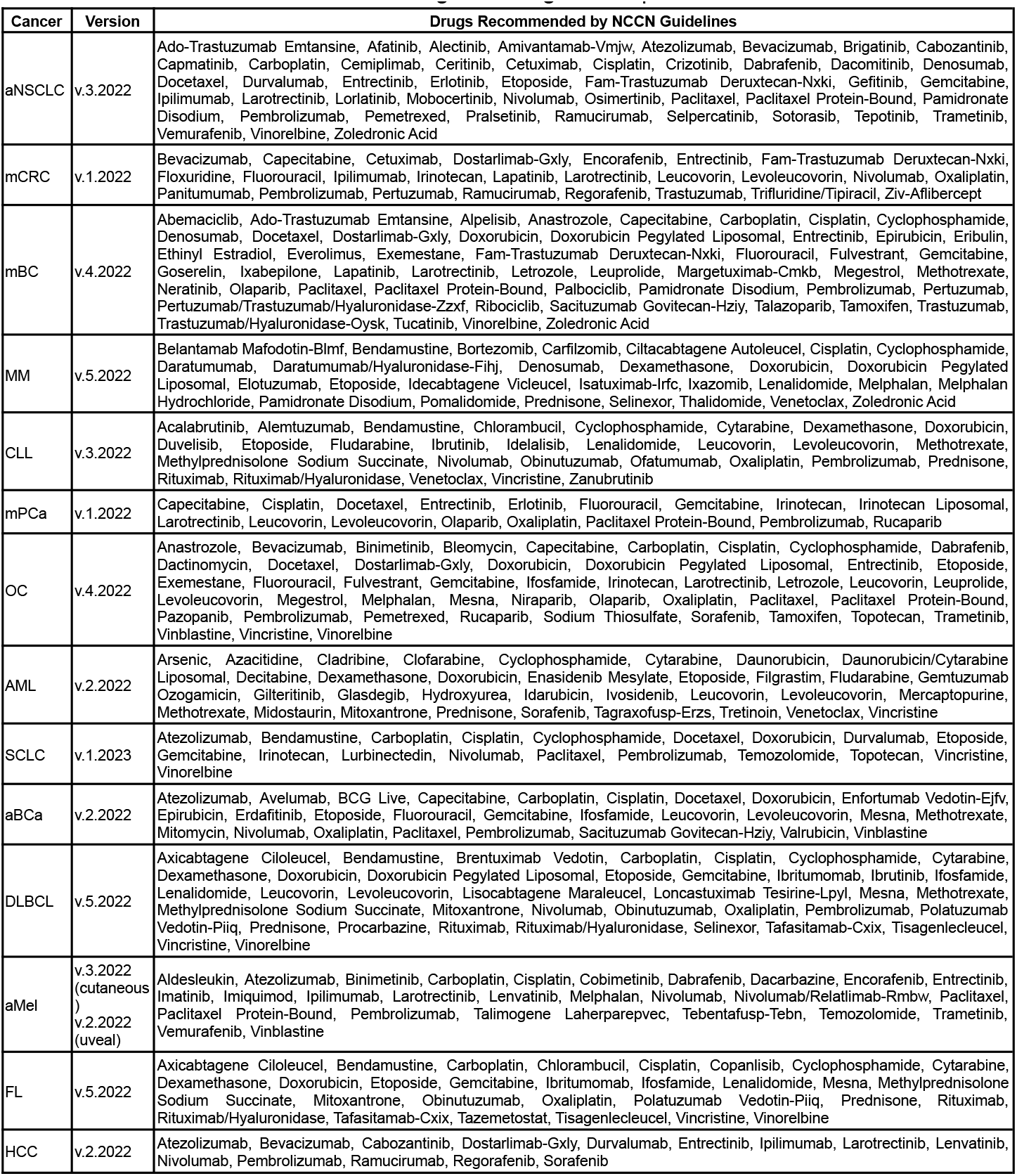
Latest available version (up to August 29, 2022) for NCCN guidelines of each cancer extracted from NCCN Drugs & Biologics Compendium.

**Supplementary Table 5.** Proportion of patients receiving off-label drugs in 14 cancers for different lines of treatments.

| Cancer | 1L | 2L | 3L | 4L | 5L | In Any Line of Treatment |
| --- | --- | --- | --- | --- | --- | --- |
| aNSCLC | 3.3% | 2.7% | 3.7% | 5.6% | 7.5% | 5.4% |
| mCRC | 0.9% | 2.6% | 3.3% | 4.3% | 6.0% | 3.6% |
| mBC | 8.8% | 10.6% | 12.9% | 15.1% | 19.3% | 25.9% |
| MM | 2.2% | 4.0% | 5.4% | 6.2% | 10.3% | 7.5% |
| CLL | 2.7% | 6.5% | 8.5% | 10.2% | 11.4% | 6.4% |
| mPCa | 5.9% | 14.3% | 19.9% | 26.0% | 25.5% | 13.8% |
| OC | 12.3% | 17.9% | 20.0% | 23.7% | 32.9% | 26.7% |
| AML | 35.0% | 44.7% | 51.8% | 57.5% | 63.0% | 53.1% |
| SCLC | 96.7% | 56.2% | 55.7% | 57.7% | 69.7% | 97.6% |
| aBCa | 39.0% | 37.8% | 46.7% | 43.9% | 43.9% | 50.1% |
| DLBCL | 27.8% | 82.9% | 83.1% | 75.5% | 83.5% | 41.2% |
| aMel | 5.0% | 13.3% | 18.2% | 25.8% | 26.2% | 11.4% |
| FL | 49.3% | 49.2% | 52.8% | 47.2% | 44.2% | 59.7% |
| HCC | 3.1% | 12.1% | 11.5% | 18.2% | 36.7% | 6.6% |
| **Whole Population** | 12.3% | 12.6% | 13.7% | 14.8% | 18.1% | 18.3% |

**Supplementary Table 6.** Proportion of patients receiving off-guideline drugs in 14 cancers for different lines of treatments.

| Cancer | 1L | 2L | 3L | 4L | 5L | In Any Line of Treatment |
| --- | --- | --- | --- | --- | --- | --- |
| aNSCLC | 1.1% | 1.8% | 2.5% | 4.3% | 5.7% | 2.5% |
| mCRC | 0.8% | 1.8% | 2.3% | 3.2% | 4.5% | 2.7% |
| mBC | 2.4% | 3.9% | 3.8% | 4.5% | 5.9% | 7.2% |
| MM | 0.5% | 1.7% | 2.9% | 2.7% | 4.3% | 3.2% |
| CLL | 2.1% | 4.6% | 6.0% | 6.8% | 9.6% | 4.6% |
| mPCa | 1.1% | 3.4% | 6.9% | 8.1% | 10.6% | 3.4% |
| OC | 0.8% | 1.5% | 2.1% | 3.1% | 3.6% | 2.9% |
| AML | 2.7% | 6.7% | 12.7% | 19.2% | 26.7% | 7.9% |
| SCLC | 0.7% | 8.5% | 9.7% | 5.7% | 7.4% | 5.5% |
| aBCa | 2.0% | 7.4% | 16.6% | 19.3% | 22.0% | 8.1% |
| DLBCL | 1.3% | 6.9% | 8.3% | 13.5% | 16.5% | 3.6% |
| aMel | 2.7% | 5.6% | 6.2% | 8.6% | 7.8% | 5.2% |
| FL | 2.3% | 8.8% | 17.4% | 24.0% | 32.7% | 6.0% |
| HCC | 2.9% | 9.7% | 9.5% | 12.5% | 23.3% | 5.4% |
| **Whole Population** | 1.3% | 3.3% | 4.4% | 5.4% | 6.9% | 3.9% |

**Supplementary Table 7.**
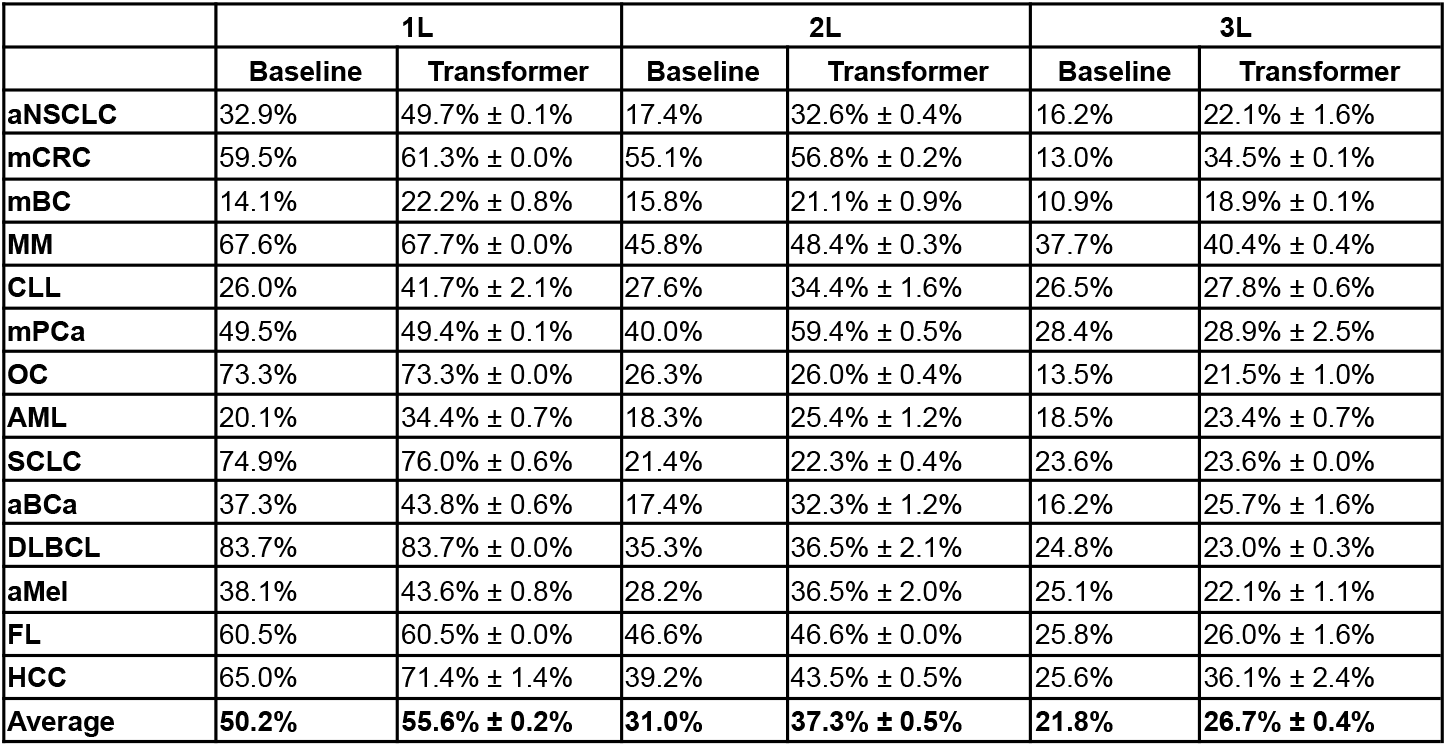
Test accuracy of predicting treatments in each line. Here we define the baseline as predicting the standard-of-care (i.e. most commonly used) drug for each cancer and line of treatment.

**Supplementary Table 8.** Test AUPRC of predicting off-guideline usage in each line. Here we compare the performance of a logistic regression model trained on 1) the original patient characteristics (LR) and 2) the patient embedding learned by the transformer.

|  | 1L |  | 2L |  | 3L |  |
| --- | --- | --- | --- | --- | --- | --- |
|  | LR | Transformer | LR | Transformer | LR | Transformer |
| **aNSCLC** | 0.065 ± 0.002 | 0.082 ± 0.003 | 0.104 ± 0.001 | 0.100 ± 0.003 | 0.117 ± 0.004 | 0.116 ± 0.004 |
| **mCRC** | 0.061 ± 0.005 | 0.063 ± 0.002 | 0.085 ± 0.005 | 0.139 ± 0.002 | 0.110 ± 0.006 | 0.129 ± 0.005 |
| **mBC** | 0.122 ± 0.006 | 0.120 ± 0.010 | 0.147 ± 0.024 | 0.218 ± 0.009 | 0.123 ± 0.009 | 0.220 ± 0.004 |
| **MM** | 0.044 ± 0.001 | 0.046 ± 0.009 | 0.069 ± 0.003 | 0.119 ± 0.008 | 0.084 ± 0.005 | 0.134 ± 0.003 |
| **CLL** | 0.084 ± 0.006 | 0.130 ± 0.001 | 0.131 ± 0.011 | 0.270 ± 0.014 | 0.137 ± 0.019 | 0.412 ± 0.039 |
| **mPCa** | 0.112 ± 0.007 | 0.187 ± 0.004 | 0.131 ± 0.014 | 0.083 ± 0.007 | 0.134 ± 0.023 | 0.213 ± 0.020 |
| **OC** | 0.054 ± 0.003 | 0.066 ± 0.006 | 0.088 ± 0.034 | 0.104 ± 0.012 | 0.106 ± 0.010 | 0.117 ± 0.019 |
| **AML** | 0.127 ± 0.006 | 0.200 ± 0.014 | 0.254 ± 0.011 | 0.313 ± 0.018 | 0.282 ± 0.014 | 0.427 ± 0.006 |
| **SCLC** | 0.054 ± 0.003 | 0.055 ± 0.002 | 0.220 ± 0.013 | 0.211 ± 0.022 | 0.138 ± 0.013 | 0.279 ± 0.020 |
| **aBCa** | 0.084 ± 0.008 | 0.087 ± 0.002 | 0.236 ± 0.012 | 0.193 ± 0.009 | 0.269 ± 0.017 | 0.343 ± 0.046 |
| **DLBCL** | 0.046 ± 0.004 | 0.049 ± 0.004 | 0.109 ± 0.016 | 0.191 ± 0.020 | 0.155 ± 0.023 | 0.377 ± 0.034 |
| **aMel** | 0.152 ± 0.008 | 0.268 ± 0.013 | 0.248 ± 0.011 | 0.146 ± 0.015 | 0.170 ± 0.025 | 0.076 ± 0.007 |
| **FL** | 0.081 ± 0.005 | 0.106 ± 0.017 | 0.177 ± 0.011 | 0.278 ± 0.012 | 0.279 ± 0.009 | 0.337 ± 0.006 |
| **HCC** | 0.086 ± 0.007 | 0.148 ± 0.009 | 0.408 ± 0.059 | 0.479 ± 0.025 | 0.248 ± 0.022 | 0.747 ± 0.004 |
| **Average** | 0.084 ± 0.001 | **0.115 ± 0.003** | 0.172 ± 0.003 | **0.203 ± 0.007** | 0.168 ± 0.005 | **0.280 ± 0.002** |

**Supplementary Table 9.**
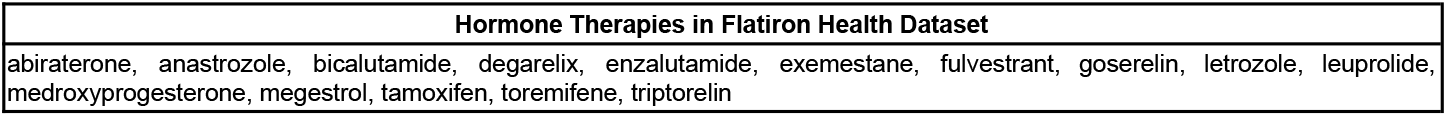
The 15 hormone therapies included in the Flatiron Health dataset.

**Supplementary Table 10.** Evaluation of hormone drugs as monotherapy in ovarian cancer for 2L+ on the FH-FMI CGDB dataset. Here the experiment arm is the patients who received the hormone drugs as monotherapy (e.g. letrozole alone).

| Drug | Number of Patients | Control | N control | HR (95% conf) | Predictive Effect of Mutation TP53 |  |  |
| --- | --- | --- | --- | --- | --- | --- | --- |
|  |  |  |  |  | Interaction HR (95% CI) | HR (95% CI) for patients with TP53 mutation | HR (95% CI) for patients without TP53 mutation |
| Letrozole | 120 | Non-hormone treatments | 2438 | 0.58 (0.44, 0.75) | 2.79 (1.43, 5.43) | 0.94 (0.72, 1.24) | 0.35 (0.19, 0.63) |
|  |  | standard of care - Doxorubicin Pegylated Liposomal | 522 | 0.47 (0.35, 0.62) | 1.90 (0.88, 4.10) | 0.69 (0.50, 0.95) | 0.39 (0.20, 0.76) |
| Tamoxifen | 108 | Non-hormone treatments | 2438 | 0.63 (0.48, 0.83) | 1.51 (0.68, 3.31) | 0.89 (0.65, 1.21) | 0.52 (0.26, 1.03) |
|  |  | standard of care - Doxorubicin Pegylated Liposomal | 522 | 0.52 (0.39, 0.68) | 0.90 (0.38, 2.14) | 0.62 (0.42, 0.90) | 0.61 (0.26, 1.44) |
| Anastrozole | 92 | Non-hormone treatments | 2438 | 0.57 (0.42, 0.76) | 1.66 (0.85, 3.24) | 0.74 (0.55, 0.99) | 0.44 (0.24, 0.80) |
|  |  | standard of care - Doxorubicin Pegylated Liposomal | 522 | 0.47 (0.34, 0.64) | 1.12 (0.51, 2.49) | 0.55 (0.39, 0.77) | 0.43 (0.21, 0.87) |

**Supplementary Table 11.** Evaluation of nivolumab in mPCa for 2L+. Here the experiment arm is the patients with mPCa who received treatments containing the nivolumab (e.g. nivolumab alone or in combination with other drugs). The control arm is the patients 1) who took treatments not containing nivolumab; 2) standard of care, defined as the most commonly used treatment. Hazard ratios are adjusted for age, sex, race, ECOG, stage, histology and molecular alterations, practice type, insurance type, year of receiving treatment.

| Control | N control | HR (95% conf) |
| --- | --- | --- |
| Treatments not containing nivolumab | 3167 | 0.93 (0.59, 1.47) |
| standard of care - Gemcitabine,Paclitaxel Protein-Bound | 1273 | 0.82 (0.51, 1.32) |

**Supplementary Table 12.** Biosimilars included in the data. Drugs and their biosimilars are grouped together in the analysis.

| Drug | Biosimilars |
| --- | --- |
| Bevacizumab | Bevacizumab-Awwb, Bevacizumab-Bvzr |
| Rituximab | Rituximab-Abbs, Rituximab-Arrx, Rituximab-Pvvr |
| Trastuzumab | Trastuzumab-Anns, Trastuzumab-Dkst, Trastuzumab-Dttb, Trastuzumab-Pkrb, Trastuzumab-Qyyp |

**Supplementary Table 13.** Biomarkers captured for each cancer available in the Flatiron Health dataset.

| Cancer | Biomarkers |
| --- | --- |
| aNSCLC | ALK, BRAF, EGFR, KRAS, PDL1, ROS1 |
| mCRC | BRAF, KRAS, MMR/MSI, NRAS |
| mBC | BRCA, ER, HER2, PDL1, PR |
| mPCa | BRCA |
| OC | BRCA, GIS, HRD |
| SCLC | PDL1 |
| aBCa | FGFR, PDL1 |
| aMel | BRAF, KIT, NRAS, PDL1 |
| HCC | PDL1 |

